# Physical function domain associations with cognitive domains in community-dwelling older adults

**DOI:** 10.64898/2026.06.29.26356840

**Authors:** Jeongwoon Kim, Bryan Monterro Herrera, Chadsley Wessinger, Brittany Armstrong, Jennifer Etnier, Kyoung Shin Park

**Affiliations:** Division of Geriatrics and Gerontology, Department of Medicine, Emory University School of Medicine, GA, USA; Department of Kinesiology, University of North Carolina – Greensboro, NC, USA

**Author notes:** Corresponding Author: Jeongwoon Kim.

**Keywords:** Older adults, physical function, cognitive domains, cognitively unimpaired, sedentary, community-dwelling

## Abstract

**Objectives:** Physical and cognitive aging do not occur uniformly, yet associations between specific physical function and cognitive domains in sedentary older adults remain unclear. This exploratory cross-sectional study examined associations between multiple physical function domains and cognitive outcomes in sedentary, community-dwelling, cognitively unimpaired older adults.

**Methods:** Fifty-eight older adults (70.7±4.7 years; 84.5% Female) completed handgrip strength, 30-second chair stand, timed up and go (TUG), brisk walk, and 6-minute walk (6MWT) assessment. Cognitive outcomes included global cognition using the Montreal Cognitive Assessment (MoCA), and working memory, episodic memory, attentional inhibition, and cognitive flexibility using the NIH Toolbox. Linear regression models adjusted for age, sex, education, body mass index, and brachial pulse pressure. False discovery rate (FDR) correction was applied.

**Results:** Greater 6MWT distance was associated with better episodic memory performance after FDR correction (*β*=0.49, *p_a_*=0.028). Additional inverse associations were observed between TUG performance and global cognition (*β*=-0.34, *p_a_*=0.166) and attentional inhibition (*β*=-0.32, *p_a_*=0.180), and between gait speed and global cognition (*β*=-0.33, *p_a_*=0.166) and episodic memory performance (*β*=-0.32, *p_a_*=0.166), however, these did not survive FDR correction. Handgrip strength and chair stand performance were not associated with cognitive outcomes.

**Conclusions:** These exploratory findings suggest that locomotor-based functional tasks may demonstrate stronger cognitive associations than strength measures in sedentary, cognitively unimpaired older adults. Tasks involving sustained locomotion and adaptive movement may place greater cognitive-motor demands, potentially increasing sensitivity to subtle cognitive variation. Larger longitudinal and multimodal studies are needed to determine whether these associations reflect reliable differential patterns across physical and cognitive domains.

**Statement of Research Significance:** *Research Question(s) or Topic(s):* Physical abilities, such as strength, walking, and endurance, and cognitive abilities, such as memory, and attention, often decline as people get older. We examined whether different physical abilities were related to different cognitive abilities in older adults who were physically inactive but not cognitive impaired.

*Main Findings:* Older adults with better walking endurance performed better on a memory test. Walking and mobility measures also showed preliminary relationships with overall cognitive ability and attention, while measures of upper- and lower-body strength were not related to cognitive abilities. However, these relationships need to be confirmed in future studies.

*Study Contributions:* Our findings suggest that not all physical and cognitive abilities are related in the same way. Walking endurance and mobility may provide information about memory in older adults for future research to identify people at risk for cognitive decline and develop more effective physical activity programs.

## Introduction

A 2023 report from the United Nations projects that the global population of adults ≥65 years will more than double by 2050, reaching 1 in 6 people worldwide (United Nations Department of Economic and Social Affairs, 2023). Age-related declines in both physical (Jackson et al., 2009) and cognitive function (Harada et al., 2013) are common in this population, contributing to loss of independence and quality of life (Fusco et al., 2012; Prasad et al., 2021).

These declines are not uniform, with both physical and cognitive functions encompassing multiple domains that deteriorate along different trajectories (Bendayan et al., 2017; Harada et al., 2013). For example, previous studies suggest that gait speed begins to decline at an earlier age than handgrip strength (Lynch et al., 2025; Ogawa et al., 2022) and that gait speed, lower body strength, and handgrip strength may follow distinct age-related patterns (Hoekstra et al., 2020). Cognitive domains show similar heterogeneity; crystallized intelligence remains relatively stable with age (Deary et al., 2009) whereas executive function and memory may begin to decline as early as midlife (Hedden & Gabrieli, 2004). Together, these findings indicate that physical and cognitive aging follow domain-specific patterns, and emerging evidence points to potentially selective associations between physical and cognitive function (Clouston et al., 2013). Clarifying these relationships could identify early markers of cognitive and functional decline and inform targeted interventions to support healthy aging.

Given the domain-specific patterns of decline, it is crucial to consider the key domains of physical function individually. These four domains—mobility, gait, muscular strength, and aerobic endurance (Dias, 2014)—depend on distinct physiological systems. Mobility and gait rely heavily on neuromotor control (Rosso et al., 2013), muscular strength depends on skeletal muscle (Goodpaster et al., 2006) and metabolic health (Yang et al., 2012), and aerobic endurance is supported by cardiorespiratory fitness (Kaminsky et al., 2019). Despite their reliance on different systems, each of these physical function domains has been previously linked to healthy aging, predicting mortality (Andrasfay, 2020; Cooper et al., 2010). Furthermore, these domains influence the brain through partially distinct pathways, resulting in domain-specific associations (Erickson et al., 2014; Erickson et al., 2009; Ezzati et al., 2015; Tian et al., 2017). Together with the domain-specific patterns of cognitive aging discussed above, this framework motivates examining whether different physical function domains demonstrate differential associations with cognitive performance.

Cognition can similarly be partitioned into different domains, with executive function and episodic memory central for maintaining independence in older adults. Executive function, composed of inhibitory control, working memory, and cognitive flexibility, supports goal-oriented behaviors and higher-order processes, such as planning (Diamond, 2013). Episodic memory enables retrieval of personal experiences and is essential for autobiographic continuity, prospective planning, and adaptive everyday functioning (Tulving, 2002). Meta-analytic and empirical evidence in older adults supports domain-specific links between physical and cognitive function: mobility and gait have been consistently associated with executive function (Demnitz et al., 2016; Desjardins-Crepeau et al., 2014; Martin et al., 2013), muscular strength has been related to both executive function and episodic memory (Wang et al., 2025; Yoon et al., 2018; Zammit et al., 2019), and aerobic endurance shows similar broad associations of benefits to both executive function and episodic memory (Oberlin et al., 2025).

Despite this evidence, limited works have examined multiple domains of physical and cognitive function in cognitively unimpaired, sedentary older adults in a single study. This population is relevant because sedentary behavior in later life has been associated with elevated risk for functional decline (Garcia Meneguci et al., 2021), frailty (Song et al., 2015), and poorer cognitive outcomes (Yan et al., 2020). Accordingly, cognitively unimpaired but sedentary older adults may represent an important target population for lifestyle interventions that emphasize specific physical function components most relevant for preserving cognitive domains at risk for accelerated decline in this population.

Accordingly, we conducted exploratory cross-sectional analyses to characterize associations between distinct physical function domains and cognitive performance in sedentary, community-dwelling, and cognitively unimpaired older adults. Given the modest sample size and cross-sectional design, these analyses were intended to be hypothesis-generating and to identify preliminary patterns rather than test definitive domain-specific hypotheses. In light of the prior evidence for domain-specific patterns of aging summarized above, we explored whether mobility- and gait-based tasks would show stronger associations with executive function, whereas muscular strength and aerobic endurance may be associated with both executive function and episodic memory. Findings are intended to inform the design and interpretation of subsequent longitudinal and intervention work that can rigorously test these domain-specific relationships.

## Methods

### Participants

Baseline data were drawn from two clinical trials (NCT06364189, NCT06496425) that were recruited from community settings in Greensboro, North Carolina using flyers and social platforms (i.e., FaceBook). Inclusion criteria included ages ≥ 65 years, the ability to speak and understand English, self-reported engagement in less than 60 min/week of moderate-to-vigorous aerobic physical activity and no resistance physical activity per week, self-report of absence of neurological or functional impairment and any health conditions preventing fitness and functional testing, and absence of cognitive impairment based upon the Montreal Cognitive Assessment (MoCA). All participants scored ≥ 21 on the MoCA—given that the original MoCA cutoff (≥26) often overclassifies older adults with limited education and minority backgrounds, 21–23 has been supported as new cutoff scores in screening community-dwelling samples without cognitive impairment (Carson et al., 2018; Milani et al., 2018; Rossetti et al., 2011).

Exclusion criteria further included presence of neurodegenerative disease, self-reported anxiety or depression, excessive alcohol use (>14 alcoholic drinks per week), drug use, and current use of cancer treatment medication.

### Study Design

Upon indicating study interest, participants were contacted by study staff to complete a series of questionnaires to determine their eligibility and provide demographic information. These online questionnaires included the ACSM Health History Questionnaire (HSQ) (Kaminsky et al., 2014) and Physical Activity Readiness Questionnaire (PAR-Q) (Thomas et al., 1992) for contraindications of exercise, the Center for Epidemiologic Studies Depression Scale Revised (CESD-R) (Van Dam & Earleywine, 2011), to assess anxiety and depression status, and the Telephone Interview for Cognitive Status (TICS) (de Jager et al., 2003) for the preliminary assessment of cognitive impairment. Eligible participants were scheduled to complete a baseline visit. During the baseline visit, participants were informed of study procedures and potential risks before providing their written consent. Participants then completed the MoCA to further determine their eligibility, before completing a battery of cognitive tests. Following this, participants completed a series of anthropometric assessments followed by a series of functional assessments. All experimental procedures were approved by the Institutional Review Board at University of North Carolina - Greensboro (IRB-FY22-119), and in accordance with the *Declaration of Helsinki* for human subject research.

### Cognitive Assessments

MoCA was administered and scored by a certified member of the study team according to the traditional maximum score of 30 as a measure of global cognitive function (Nasreddine et al., 2005). As a part of the NIH Toolbox© (NIH-TB; Version 2), participants completed the list sort working memory (LSWM) task to assess working memory, the picture sequence memory (PSM) task to assess episodic memory, the flanker task to assess attentional inhibition, and the dimensional change card sort (DCCS) task to assess cognitive flexibility on an iPad (Weintraub et al., 2013). Performance on each task was assessed using the uncorrected score that scales performance to a standard score with a mean of 100 and a standard deviation of 15 that does not adjust for the participant’s age, education, or other characteristics and can be compared to the average individual in the United States (Casaletto et al., 2015).

### Anthropometric Assessment

Following the cognitive assessments, participants completed height (SECA, Hamburg, Germany) and weight (Etekcity, Anaheim, CA, USA) assessments to calculate body mass index (BMI), and blood pressure assessments to measure systolic and diastolic blood pressure (Omron, Kyoto, Japan). Pulse pressure (PP) was calculated from these blood pressure measurements as the difference between systolic and diastolic blood pressure.

### Functional Assessment

Following the anthropometric assessments, participants completed a series of functional assessments including handgrip strength, 30-second chair stand test, the timed-up and go (TUG), 10-meter walk test, and the 6-minute walk test (6MWT) to assess different physical functional domains. Upper body strength was measured through handgrip strength dynamometry (Scandidact, Denmark) of the dominant hand averaged across 3 repetitions (Reijnierse et al., 2017). Lower body strength was measured through the number of successful repetitions during the 30-second chair stand test (Jones et al., 1999). Mobility was assessed through the time required to complete the TUG (van Iersel et al., 2008). Gait was assessed through the time required to travel from 2-meter to 8-meter during the 10-meter walk test at a brisk pace (Graham et al., 2008). Aerobic endurance was assessed using the total distance traveled during the 6MWT (Dourado et al., 2021).

### Statistical Analyses

While 62 participants provided consent and were enrolled in the study, 4 participants were excluded from these analyses due to missing cognitive and/or physical function data (*n*=58). Associations between physical function and cognitive outcomes were examined through hierarchical linear regressions after adjusting for potential confounding variables using SPSS (v31, Armonk, NY, USA) and R (v4.5.1) with an *α*-level of *p* <0.05 to indicate statistical significance. Age, sex, years of education, BMI, and PP were included in model 1 as *a priori* covariates. Meta-analytic evidence suggests that age (Verhaeghen & Salthouse, 1997), sex (Levine et al., 2021), education (Noble et al., 2021), BMI (Qu et al., 2020), and PP (Cecelja et al., 2025) influence cognition in older adults. For model 2, physical function outcomes were included as predictors in separate models to evaluate the associations with specific cognitive domains. Sensitivity analysis indicated that the present sample provided 80% power to detect effect sizes *f^2^*=0.15 or larger for models including 5 covariates and 1 predictor at *α*=0.05. Overall fit of the model was assessed through using the omnibus ANOVA *F*-statistic, *R^2^,* and adjusted *R^2^*. The incremental contribution of each physical function measure was evaluated using the change in the ANOVA and *R^2^* to represent the additional variation explained in each model. Regression results were evaluated through unstandardized coefficient (B) with the corresponding 95% confidence interval (95% CI), standardized regression coefficient (β), and exact p-values.

Univariate normality was evaluated using skewness, kurtosis, and visually confirmed through histograms and P-P plots. One outlier for handgrip strength, TUG, and gait (≥3 standard deviations from mean) was winsorized with the corresponding highest non-outlier value to preserve the distribution while reducing the influence of extreme observations. Following winsorization, all cognitive and physical function variables demonstrated acceptable distributional properties (skew < 1.03, kurtosis < 1.85) supporting the use of parametric regression. To account for multiple comparisons, false discovery rate (FDR) correction was applied using the Benjamini-Hochberg correction for all associations between physical function and cognitive performance with unadjusted *p* values and adjusted *p* values (*p_a_*) reported throughout. Associations surviving FDR correction were interpreted as the strongest statistical evidence, whereas significant uncorrected associations were interpreted as exploratory signals requiring confirmation. As the majority of our sample (*n*=49) were female, we conducted *post-hoc* sensitivity analyses excluding males.

## Results

### Demographics

58 older adults (70.7±4.7 years; 84.5% Female) were included in these analyses. For this sample, the average years of education was 15.9±2.2 years, average BMI was 32.5±6.1 kg/m^2^, and average PP was 49.1±18.3 mmHg. (Table 1)

**Table 1.** Descriptive Statistics of Study Population

| N = 58 |  |
| --- | --- |
| Demographics |  |
| Age (years) | 70.7 ± 4.7 |
| Sex (% Female) | 84.5% |
| BMI (kg/m<sup>2</sup>) | 32.5 ± 6.1 |
| Education (years) | 15.9 ± 2.2 |
| Systolic Blood Pressure (mmHg) | 124.5 ± 19.9 |
| Diastolic Blood Pressure (mmHg) | 75.4 ± 9.1 |
| Pulse Pressure (mmHg) | 49.1 ± 18.3 |
| Cognitive Measures |  |
| MoCA | 26.6 ± 1.9 |
| List Sort Working Memory* | 99.4 ± 8.3 |
| Picture Sequence Memory* | 98.2 ± 10.1 |
| Flanker Inhibitory Control* | 94.5 ± 6.8 |
| Dimensional Change Card Sort* | 101.8 ± 7.5 |
| Functional Outcomes |  |
| Handgrip Strength (kg) | 24.2 ± 5.9 |
| 30-Second Chair Stand (#) | 10.1 ± 2.8 |
| Timed Up and Go (s) | 8.9 ± 1.5 |
| 10-Meter Walk (s) | 4.0 ± 0.6 |
| 6-Minute Walk Test (m) | 428.8 ± 75.2 |

Note: Values presented as mean ± SD unless specified otherwise; \* - indicates that values are uncorrected standardized scores; BMI – body mass index; MoCA – Montreal Cognitive Assessment

### Regression Analyses (Covariates)

In Model 1, the inclusion of the covariates, age, sex, education, BMI, and PP, did not predict MoCA scores (*F(5,52)=*1.66; *R^2^*=0.14; adjusted *R^2^=*0.05; *p*=0.162), LSWM (*F(5,52)=*1.97; *R^2^*=0.16; Adjusted *R^2^=*0.08; *p*=0.099), flanker (*F(5,52)=*0.76; *R^2^*=0.07; adjusted *R^2^=-*0.02; *p*=0.583) or DCCS performance (*F(5,52)=*1.44; *R^2^*=0.12; adjusted *R^2^=*0.04; *p*=0.227).

These covariates significantly predicted PSM performance (*F(5,52)=*2.77; *R^2^*=0.21; adjusted *R^2^=*0.14; *p*=0.027). A summary of these analyses is presented in Table 2. Upon excluding male participants, the covariates did not predict MoCA scores (*F(4,44)=*1.55; *R^2^*=0.12; adjusted *R^2^=*0.04; *p*=0.203), LSWM (*F(4,44)=*1.58; *R^2^*=0.13; Adjusted *R^2^=*0.05; *p*=0.198), PSM (*F(4,44)=*2.42; *R^2^*=0.18; adjusted *R^2^=*0.11; *p*=0.063), flanker (*F(4,44)=*0.81; *R^2^*=0.07; adjusted *R^2^=-*0.02; *p*=0.523) or DCCS performance (*F(4,44)=*1.39; *R^2^*=0.11; adjusted *R^2^=*0.03; *p*=0.252). A summary of these analyses is presented in Supplementary Table 1.

**Table 2.** Influence of Potential Covariates on Cognitive Measures

|  | MoCA | LSWM* | PSM* | Flanker* | DCCS* |
| --- | --- | --- | --- | --- | --- |
| Model ($F(5,52)$, $R^2$, Adj. $R^2$, $p$) | 1.66, 0.14, 0.05, 0.162 | 1.97, 0.16, 0.08, 0.099 | 2.77, 0.21, 0.14, 0.027 | 0.76, 0.07, -0.02 0.583 | 1.44, 0.12, 0.04, 0.227 |
| Age ($\beta$, $p$) | -0.14, 0.273 | -0.16, 0.234 | **-0.40, 0.003** | -0.19, 0.164 | **-0.30, 0.029** |
| Sex ($\beta$, $p$) | -0.08, 0.546 | -0.20, 0.130 | -0.04, 0.779 | 0.03, 0.807 | 0.02, 0.861 |
| Education ($\beta$, $p$) | 0.23, 0.088 | **0.27, 0.046** | 0.16, 0.201 | -0.15, 0.285 | -0.15, 0.258 |
| BMI ($\beta$, $p$) | 0.22, 0.110 | 0.05, 0.698 | 0.10, 0.425 | -0.08, 0.543 | -0.14, 0.302 |
| PP ($\beta$, $p$) | 0.15, 0.256 | 0.14, 0.285 | -0.08, 0.530 | 0.13, 0.356 | -0.03, 0.799 |

Note: \* - indicates that values are uncorrected standardized scores; Adj. $R^2$ – Adjusted $R^2$ ; BMI – body mass index; PP – pulse pressure; MoCA – Montreal Cognitive Assessment; LSWM – List Sort Working Memory; PSM – Picture Sequence Memory; DCCS – Dimensional Change Card Sort

### Upper Body Strength

Handgrip strength did not improve the model for predicting MoCA (*ΔF(1,51)=*0.40; *ΔR^2^*=0.01; *β*=-0.11; *p*=0.532; *p_a_*=0.782), LSWM (*ΔF(1,51)=*0.56; *ΔR^2^*=0.01; *β*=-0.13; *p*=0.458; *p_a_*=0.763), PSM (*ΔF(1,51)=*0.25; *ΔR^2^*=0.00; *β*=-0.09; *p*=0.623; *p_a_*=0.792), flanker (*ΔF(1,51)=*0.17; *ΔR^2^*=0.00; *β*=0.08; *p*=0.685; *p_a_*=0.792), or DCCS (*ΔF(1,51)=*2.74; *ΔR^2^*=0.05; *β*=0.30; *p*=0.104; *p_a_*=0.288) performance. Similarly, upon excluding male participants, handgrip strength did not improve the models for MoCA scores (*ΔF(1,43)=*0.26; *ΔR^2^*=0.01; *β*=-0.08; *p*=0.611; *p_a_*=0.728), LSWM (*ΔF(1,43)=*0.75; *ΔR^2^*=0.02; *β*=-0.13; *p*=0.393; *p_a_*=0.614), PSM (*ΔF(1,43)=*0.17; *ΔR^2^*=0.00; *β*=0.06; *p*=0.681; *p_a_*=0.745), flanker (*ΔF(1,43)=*0.13; *ΔR^2^*=0.00; *β*=0.06; *p*=0.718; *p_a_*=0.748), DCCS (*ΔF(1,43)=*0.77; *ΔR^2^*=0.02; *β*=0.13; *p*=0.384; *p_a_*=0.614) performance.

### Lower Body Strength

Repetitions during the 30-second chair stand did not improve the model for predicting MoCA scores (*ΔF(1,51)=*1.36; *ΔR^2^*=0.02; *β*=0.17; *p*=0.249; *p_a_*=0.519), LSWM (*ΔF(1,51)=*0.12; *ΔR^2^*=0.00; *β*=0.05; *p*=0.727; *p_a_*=0.792), PSM (*ΔF(1,51)=*0.03; *ΔR^2^*=0.00; *β*=0.02; *p*=0.872; *p_a_*=0.872), flanker (*ΔF(1,51)=*2.14; *ΔR^2^*=0.04; *β*=0.22; *p*=0.150; *p_a_*=0.374), or DCCS (*ΔF(1,51)=*0.12; *ΔR^2^*=0.00; *β*=0.05; *p*=0.729; *p_a_*=0.792) performance. Upon excluding male participants, repetitions during the 30-second chair stand did not improve the model for MoCA scores (*ΔF(1,43)=*0.54; *ΔR^2^*=0.01; *β*=0.12; *p*=0.466; *p_a_*=0.650), LSWM (*ΔF(1,43)=*0.48; *ΔR^2^*=0.01; *β*=0.11; *p*=0.494; *p_a_*=0.650), PSM (*ΔF(1,43)=*0.31; *ΔR^2^*=0.01; *β*=0.09; *p*=0.579; *p_a_*=0.724), flanker (*ΔF(1,43)=*0.51; *ΔR^2^*=0.01; *β*=0.12; *p*=0.477; *p_a_*=0.650), DCCS (*ΔF(1,43)=*0.09; *ΔR^2^*=0.00; *β*=0.05; *p*=0.769; *p_a_*=0.769) performance

### Mobility

TUG times were inversely associated with MoCA scores (*ΔF(1,51)=*5.12; *ΔR^2^*=0.08; *β*=-0.34; *p*=0.028; *p_a_*=0.166) and flanker performance (*ΔF(1,51)=*4.30; *ΔR^2^*=0.07; *β*=-0.32; *p*=0.043; *p_a_*=0.180). (Figure 1A, 1B) However, these associations were not significant after FDR correction. No associations were observed for LSWM (*ΔF(1,51)=*1.21; *ΔR^2^*=0.02; *β*=-0.17; *p*=0.277; *p_a_*=0.533), PSM (*ΔF(1,51)=*0.07; *ΔR^2^*=0.00; *β*=-0.04; *p*=0.786; *p_a_*=0.819), or DCCS (*ΔF(1,51)=*0.45; *ΔR^2^*=0.01; *β*=-0.11; *p*=0.504; *p_a_*=0.782) performance. Upon excluding male participants, the MoCA scores (*ΔF(1,43)=*3.21; *ΔR^2^*=0.06; *β*=-0.32; *p*=0.080; *p_a_*=0.334) and flanker performance (*ΔF(1,43)=*2.10; *ΔR^2^*=0.04; *β*=-0.27; *p*=0.155; *p_a_*=0.481) associations were no longer significant before or after FDR correction. While an inverse association between TUG times and LSWM performance (*ΔF(1,43)=*5.85; *ΔR^2^*=0.11; *β*=-0.42; *p*=0.020; *p_a_*=0.166) emerged, this association was not significant after FDR correction. No significant associations were observed for PSM (*ΔF(1,43)=*0.17; *ΔR^2^*=0.00; *β*=-0.07; *p*=0.685; *p_a_*=0.745) or DCCS (*ΔF(1,43)=*1.55; *ΔR^2^*=0.03; *β*=-0.23; *p*=0.221; *p_a_*=0.521) performance.

**Figure 1.**
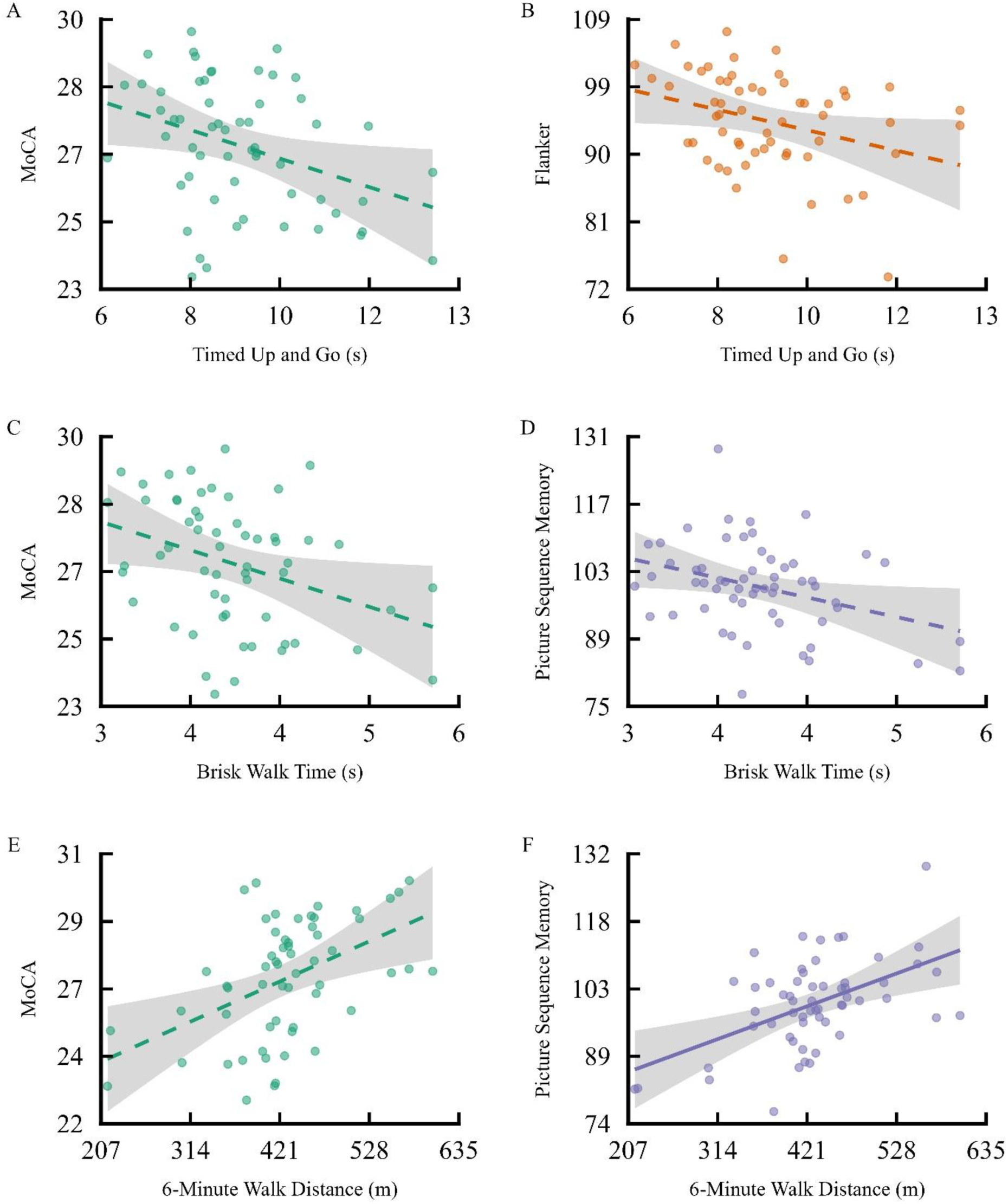
Covariate-adjusted associations of A) TUG with MoCA, B) TUG with Flanker performance, C) brisk walk time with MoCA, D) brisk walk time with PSM, E) 6MWT distance with MoCA, F) 6MWT distance with PSM. Lines represent adjusted regression estimates and shaded regions indicate 95% confidence intervals. Points represent partial residuals. Dashed lines represent associations that did not survive FDR correction

### Gait

Brisk walk times were inversely associated with MoCA scores (*ΔF(1,51)=*4.86; *ΔR^2^*=0.08; *β*=-0.33; *p*=0.032; *p_a_*=0.166) and PSM performance (*ΔF(1,51)=*4.79; *ΔR^2^*=0.07; *β*=- 0.32; *p*=0.033; *p_a_*=0.166) but were not significant following FDR correction (Figure 1C, 1D). Brisk walk times were not associated with LSWM (*ΔF(1,51)=*0.14; *ΔR^2^*=0.00; *β*=-0.06; *p*=0.714; *p_a_*=0.792), flanker (*ΔF(1,51)=*3.21; *ΔR^2^*=0.06; *β*=-0.29; *p*=0.079; *p_a_*=0.247), or DCCS (*ΔF(1,51)=*1.39; *ΔR^2^*=0.02; *β*=-0.19; *p*=0.245; *p_a_*=0.519) performance. Upon excluding male participants, the associations with MoCA scores (*ΔF(1,43)=*4.12; *ΔR^2^*=0.08; *β*=-0.34; *p*=0.049; *p_a_*=0.243) and PSM (*ΔF(1,43)=*4.70; *ΔR^2^*=0.08; *β*=-0.35; *p*=0.036; *p_a_*=0.223) performance were not significant following FDR correction. LSWM (*ΔF(1,43)=*1.17; *ΔR^2^*=0.02; *β*=-0.19; *p*=0.285; *p_a_*=0.548), flanker (*ΔF(1,43)=*2.77; *ΔR^2^*=0.06; *β*=-0.29; *p*=0.104; *p_a_*=0.370), and DCCS (*ΔF(1,43)=*1.21; *ΔR^2^*=0.02; *β*=-0.19; *p*=0.277; *p_a_*=0.548) were not associated with brisk walk times.

### Aerobic Endurance

While the 6MWT distance association with MoCA scores (*ΔF(1,51)=*9.78; *ΔR^2^*=0.14; *β*=0.47; *p*=0.003; *p_a_*=0.068) did not remain significant after correction, the positive association between 6MWT distance and PSM (*ΔF(1,51)=*11.97; *ΔR^2^*=0.15; B=0.07, 95%CI [0.03, 0.10], *β*=0.49; *p*=0.001; *p_a_*=0.028) performance survived FDR correction (Figure 1E, 1F). LSWM (*ΔF(1,51)=*1.08; *ΔR^2^*=0.02; *β*=0.17; *p*=0.304; *p_a_*=0.542), flanker (*ΔF(1,51)=*3.79; *ΔR^2^*=0.06; *β*=0.32; *p*=0.057; *p_a_*=0.204) or DCCS (*ΔF(1,51)=*0.14; *ΔR^2^*=0.00; *β*=0.06; *p*=0.715; *p_a_*=0.792) performance were not associated with 6MWT distance. After excluding males, the significant association with PSM (*ΔF(1,43)=*18.42; *ΔR^2^*=0.25; B =0.09, 95%CI [0.05, 0.13], *β*=0.60; *p*<0.001; *p_a_*=0.003) performance persisted. Associations with MoCA scores (*ΔF(1,43)=*8.59; *ΔR^2^*=0.15; *β*=0.46; *p*=0.005; *p_a_*=0.068), LSWM (*ΔF(1,43)=*1.49; *ΔR^2^*=0.03; *β*=0.21; *p*=0.229; *p_a_*=0.521), flanker (*ΔF(1,43)=*1.92; *ΔR^2^*=0.04; *β*=0.24; *p*=0.173; *p_a_*=0.481), and DCCS (*ΔF(1,43)=*0.98; *ΔR^2^*=0.02; *β*=0.17; *p*=0.327; *p_a_*=0.548) were not significant.

A detailed summary of these analyses is presented in Table 3 and a summary following the exclusion of males presented in Supplementary Table 2.

**Table 3.**
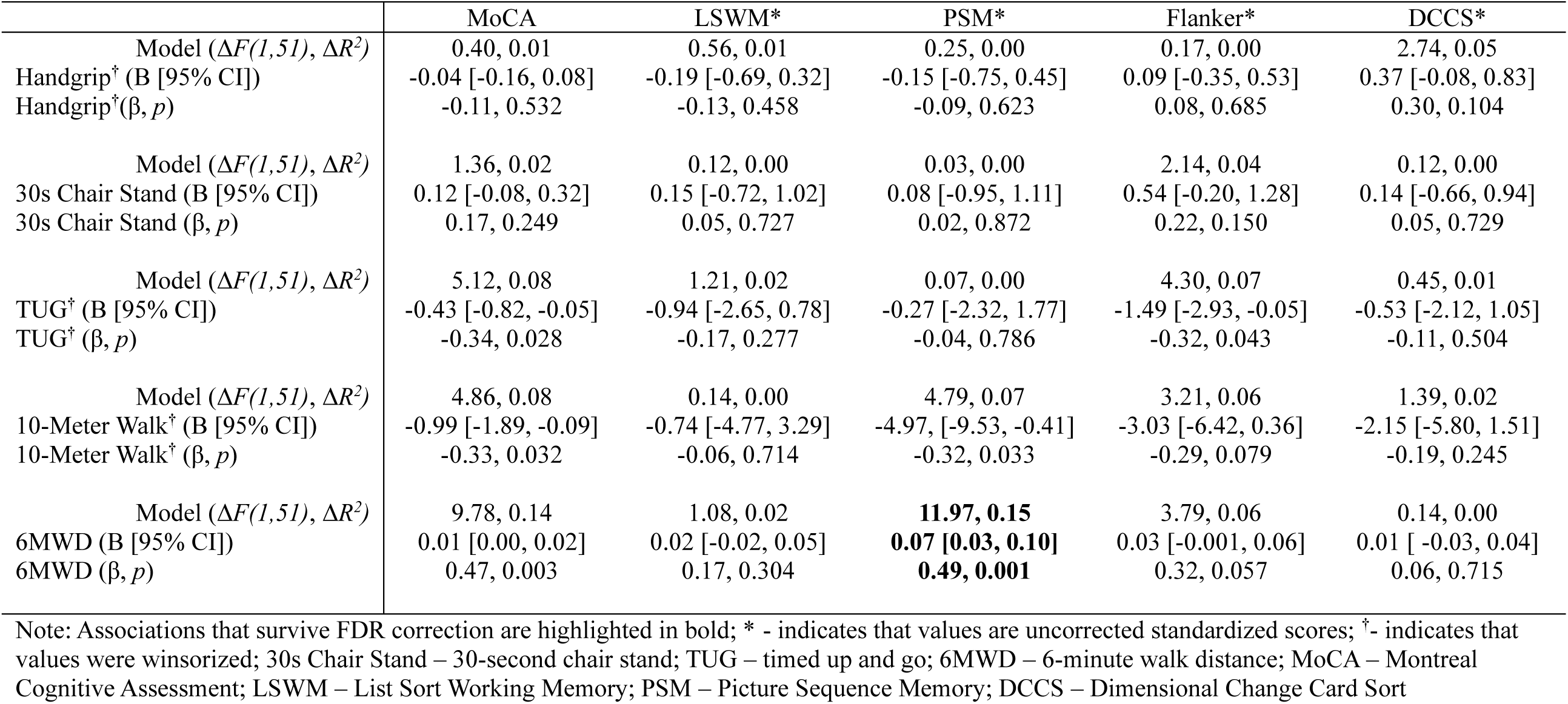
Summary of Regression Analyses Examining the Association Between Physical Function and Cognition

## Discussion

This exploratory cross-sectional study examined associations between multiple physical function domains and cognitive domains in sedentary, community-dwelling, cognitively unimpaired older adults. Although exploratory, we anticipated that mobility- and gait-based tasks would relate most strongly to executive function, whereas muscular strength and aerobic endurance might relate to both executive function and episodic memory. The clearest finding was that greater aerobic endurance, measured by 6MWT distance, was associated with better episodic memory performance after FDR correction. Additional nominal associations were observed between slower TUG and gait performance with lower global cognition and selected cognitive domains. Upper- (i.e., handgrip strength) and lower-body (i.e., 30-second chair stand) strength were not independently associated with cognitive outcomes. Taken together, these findings suggest that physical function domains may not be uniformly related to cognitive performance in sedentary cognitively unimpaired older adults and provide limited preliminary support for domain-specific physical–cognitive associations, though the evidence for domain specificity remains exploratory.

### Differential Associations Between Physical Function and Cognition

The observed pattern of associations raises the possibility that functional tasks involving locomotion, balance, endurance, and coordinated movement may demonstrate stronger cognitive associations than isolated strength measures. One potential explanation is that these tasks place greater demands on attentional allocation, sensorimotor integration, and adaptive behavioral regulation across time, potentially increasing sensitivity to subtle cognitive variation in cognitively unimpaired sedentary older adults (Donoghue et al., 2012). In contrast, isolated strength measures may reflect narrower physiological capacities that are less dependent on higher-order cognition. However, given the limited statistical power of the present study, particularly for muscular strength outcomes, these interpretations should be considered preliminary and require replication in adequately powered samples. Alternatively, the stronger association observed for the 6MWT may reflect broader contributions of aerobic endurance to cognitive function in sedentary older adults.

### Locomotor Function and Cognitive Performance

The nominal associations for TUG times and gait speed observed in this study are directionally consistent with prior studies reporting that poorer mobility and slower gait speed are associated with lower cognitive performance across multiple domains (Daimiel et al., 2020; Demnitz et al., 2016; Lundberg et al., 2024). Hartley et al. reported that slower TUG times predicted greater decline in global cognition and executive function across 4 years in 3675 middle-aged and older adults (Hartley et al., 2022), while Su et al. observed that slower gait speed was associated with poorer global cognition, memory, and executive function over 5.7 years in 459 community-dwelling older adults (Su et al., 2025). Tasks such as the TUG and brisk walking may require greater coordination of attentional focus, executive control, and sensorimotor regulation during movement (Yogev-Seligmann et al., 2008), potentially contributing to their observed associations with cognition. However, because these associations did not survive correction for multiple comparisons, they should be interpreted cautiously and viewed as preliminary patterns requiring replication. Importantly, the association between aerobic endurance and episodic memory may be particularly relevant given that episodic memory decline represents a core feature of age-related cognitive impairment and Alzheimer’s disease (Tromp et al., 2015).

Emerging evidence suggests that the relationship between locomotion and cognition may be bidirectional (Su et al., 2025). Rather than reflecting strict one-to-one correspondence between specific physical and cognitive domains, the present findings raise the possibility that shared underlying mechanisms may contribute to associations across multiple physical and cognitive outcomes. Prior work has implicated vascular dysfunction (Inoue et al., 2023), white matter hyperintensities (Murray et al., 2010), hippocampal atrophy (Rosso et al., 2017), and broader neural integrity as potential contributors to both mobility decline and cognitive impairment (Boutzoukas et al., 2021). However, these mechanisms were not directly assessed in the present study, and future longitudinal studies incorporating neuroimaging and vascular assessments will be necessary to determine whether shared neural and physiological pathways contribute to locomotor-cognitive associations in sedentary older adults.

### Muscular Strength and Cognitive Outcomes

Although muscular strength has been linked to cognitive performance in prior studies (Wang et al., 2025; Yoon et al., 2018; Zammit et al., 2019), no independent associations were observed for upper- or lower-body strength and cognitive outcomes in the present study. This contrasts with cross-sectional evidence indicating that handgrip strength is associated with multiple cognitive domains including episodic memory, working memory, and global cognition (Lundberg et al., 2024). However, previous work also suggests that sex may moderate the relationship between grip strength and cognitive performance such that a positive association between grip strength and cognitive performance is detectable in older men but not women (Prokopidis et al., 2023). Notably, females comprised 84.5% of the present sample, and the limited representation of males may have reduced our ability to detect associations between strength and cognition as previously reported in less homogeneous samples. Taken together, while muscular strength has been shown to be an important predictor of cognitive outcomes in previous studies (Lundberg et al., 2024; Prokopidis et al., 2023), these associations may be attenuated in homogeneous samples, particularly when contrasted with mobility- and endurance-based functional measures, though whether differences in cognitive-motor demands contribute to these patterns remains speculative and requires further investigation.

### Hypotheses and Future Directions

Although the present findings are exploratory and primarily cross-sectional, the observed pattern of associations raises some testable hypotheses regarding the cognitive-motor demands embedded within different functional tasks. First, it is hypothesized that functional tasks involving sustained locomotion and adaptive movement regulation may exhibit stronger cognitive associations than isolated strength measures because of their greater cognitive-motor demands. Increasingly complex locomotor tasks may differentially recruit executive and sensorimotor neural systems, thereby increasing sensitivity to subtle cognitive variation in cognitively unimpaired older adults. Future neuroimaging studies examining neural activation and connectivity during functional task performance may help clarify the cognitive-motor mechanisms underlying these relationships.

Second, it is possible that locomotor-related functional assessments may detect subtle neurocognitive vulnerability prior to the overt expression of cognitive impairment. Given the growing evidence linking vascular dysfunction (Alvarez-Bueno et al., 2020; Pase et al., 2012) and mobility impairments (Beauchet et al., 2016; de Oliveira Silva et al., 2019) to elevated risk for cognitive decline and dementia, these assessments may capture early neural inefficiencies not readily expressed through traditional cognitive screening. Future multimodal studies integrating neuroimaging, vascular markers, and sensor-derived movement metrics in longitudinal designs are necessary to determine whether baseline associations predict subsequent cognitive decline in this population and reflect early preclinical vulnerability.

Third, if replicated longitudinally, the present findings suggest that the cognitive demands embedded within physical activity interventions may influence their neurocognitive benefits.

Specifically, physical activity interventions combining executive function demands (e.g., dual-task) (Wollesen et al., 2020), complex movement patterns (e.g., dance), or continuous adaptation (e.g., open skill exercises) (Gu et al., 2019) may preferentially engage cognitive-motor systems involved in executive and memory processes (Singh et al., 2025) and provide distinct neurocognitive effects compared to interventions focused primarily on muscular or cardiovascular improvements. Future interventions directly comparing these different approaches while incorporating neuroimaging, vascular, and sensor-based assessments may help determine whether intervention-related cognitive benefits differ according to the cognitive-motor demands of physical activity itself.

### Limitations

This study has limitations that are important to note. The cross-sectional design prohibits causal inference and limits the ability to determine if declines in physical function caused cognitive changes. The study may have been underpowered to detect small to moderate associations, especially for muscular strength and some executive functions; therefore, caution is needed when interpreting results, as they could reflect limited power rather than a true absence of association. Additionally, most mobility- and gait-related associations did not survive correction for multiple comparisons and therefore should be interpreted as exploratory signals rather than robust evidence of association, informing future work designed to precisely examine domain-specific relationships. Also, the sample was mostly female, white, and highly educated, which limits how well the results apply to other groups such as males and underrepresented races and ethnicities. Although several important factors were included as covariates, other factors like sleep quality, diet, medication use, and lifetime physical activity could influence physical and cognitive performance.

These limitations are balanced by several key strengths, including the concurrent assessment of multiple physical and cognitive domains. Examining TUG, gait, 6MWT, and muscular strength alongside global cognition, executive subdomains, and episodic memory, allowed for broad assessment of potentially differential associations between physical and cognitive function across multiple physical function and cognitive measures. Another strength is the focus on sedentary yet cognitively unimpaired older adults, a population that is directly relevant for early detection and prevention prior to overt cognitive impairment. Finally, the use of well-validated physical (TUG, gait speed, 6MWT, handgrip, 30-second chair stand) and cognitive (MoCA, NIH Toolbox) assessments supports high measurement reliability.

### Conclusions

The findings from this cross-sectional study indicate that in sedentary, community-dwelling, cognitively unimpaired older adults, aerobic endurance exhibited the strongest relationship with cognition, with 6MWT distances positively associated with episodic memory performance after correcting for multiple comparisons. Mobility and gait measures exhibited additional nominal associations with global cognition and select cognitive domains, but did not survive correction and should be interpreted as preliminary. In contrast, isolated upper- and lower-body strength measures were not associated with cognitive outcomes. These exploratory findings suggest that functional tasks involving endurance and complex locomotor demands may demonstrate stronger associations with cognitive performance compared to isolated strength measures, but larger longitudinal studies are necessary to determine whether these patterns reflect reliable domain-specific patterns. Collectively, these results underscore the importance of considering the qualitative demands of functional tasks when examining physical-cognitive relationships. Simple, low-cost functional assessments such as the TUG, gait speed, and 6MWT may provide clinically meaningful information regarding cognitive vulnerability in sedentary older adults if replicated in larger prospective samples.

## Data Availability

Data Availability: The data underlying this article may be shared upon reasonable request

## Acknowledgements

Conflict of Interest: The authors have no conflict of interest to report

Sources of Support: Research reported in this presentation was supported by the National Institute on Aging of the National Institutes of Health (Award Number R61/R33 AG084479) and Alzheimer’s Association (Award Number AARF-23-1145107).

---

**Supplementary Table 1.** Influence of Potential Covariates on Cognitive Measures Excluding Males

|  | MoCA | LSWM* | PSM* | Flanker* | DCCS* |
| --- | --- | --- | --- | --- | --- |
| Model ($F(4,44)$, $R^2$, Adj. $R^2$, $p$) | 1.55, 0.12, 0.04, 0.203 | 1.58, 0.13, 0.05 0.198 | 2.42, 0.18, 0.11, 0.063 | 0.81, 0.07, -0.02, 0.523 | 1.39, 0.11, 0.03, 0.252 |
| Age ($\beta$, $p$) | -0.14, 0.335 | -0.04, 0.786 | **-0.38, 0.010** | -0.10, 0.494 | -0.26, 0.083 |
| Education ($\beta$, $p$) | 0.22, 0.139 | **0.31, 0.041** | 0.15, 0.292 | -0.15, 0.320 | -0.14, 0.337 |
| BMI ($\beta$, $p$) | 0.18, 0.233 | -0.05, 0.728 | 0.08, 0.284 | -0.16, 0.290 | -0.11, 0.456 |
| PP ($\beta$, $p$) | 0.19, 0.193 | 0.14, 0.345 | -0.04, 0.755 | 0.20, 0.190 | -0.10, 0.511 |

Note: \* - indicates that values are uncorrected standardized scores; Adj. $R^2$ – Adjusted $R^2$ ; BMI – body mass index; PP – pulse pressure; MoCA – Montreal Cognitive Assessment; LSWM – List Sort Working Memory; PSM – Picture Sequence Memory; DCCS – Dimensional Change Card Sort

**Supplementary Table 2.**
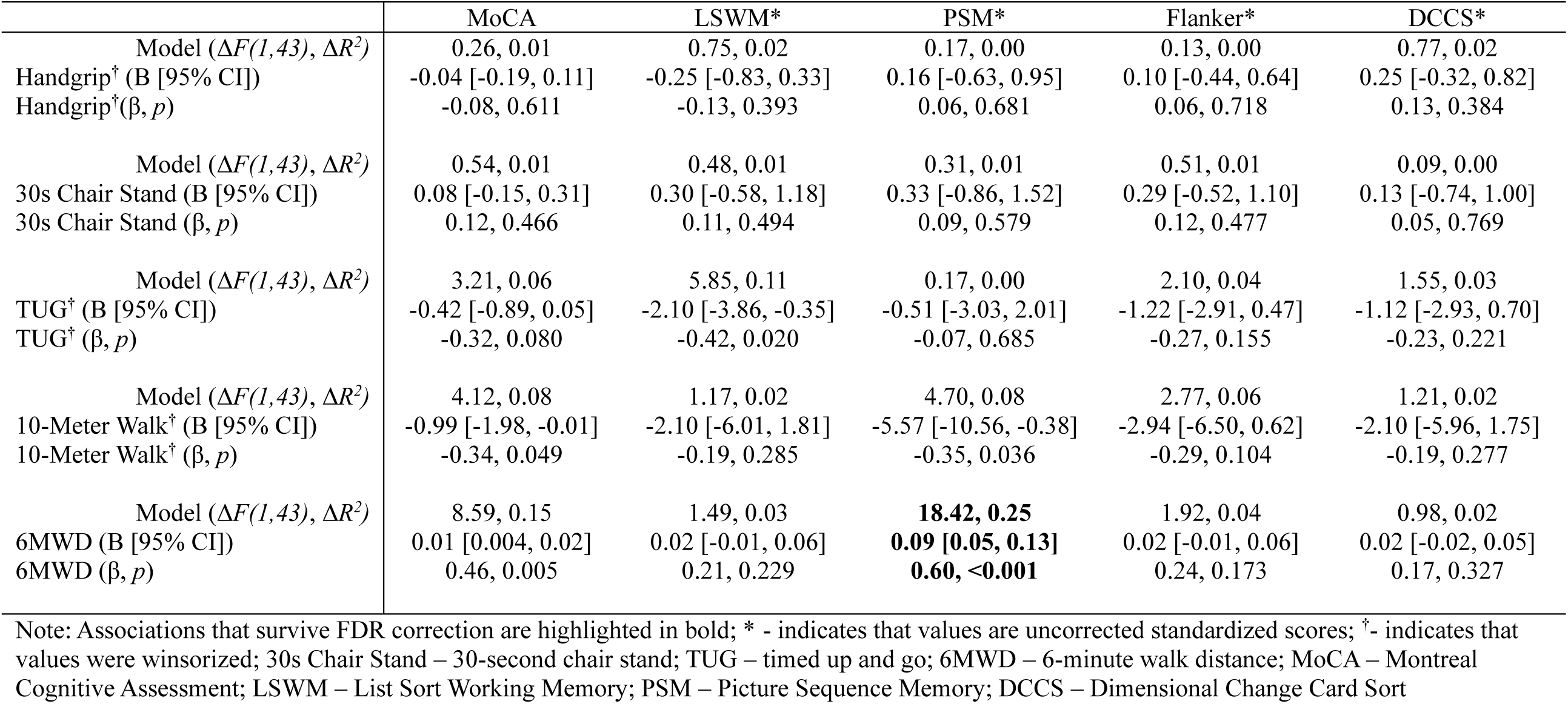
Summary of Regression Analyses Examining the Association Between Physical Function and Cognition Excluding Males

## References

1. Alvarez-Bueno, C., Cunha, P. G., Martinez-Vizcaino, V., Pozuelo-Carrascosa, D. P., Visier-Alfonso, M. E., Jimenez-Lopez, E., & Cavero-Redondo, I. (2020). Arterial Stiffness and Cognition Among Adults: A Systematic Review and Meta-Analysis of Observational and Longitudinal Studies. J Am Heart Assoc, 9(5), e014621. 10.1161/JAHA.119.014621

2. Andrasfay, T. (2020). Changes in Physical Functioning as Short-Term Predictors of Mortality. J Gerontol B Psychol Sci Soc Sci, 75(3), 630–639. 10.1093/geronb/gby133

3. Beauchet, O., Annweiler, C., Callisaya, M. L., De Cock, A. M., Helbostad, J. L., Kressig, R. W., Srikanth, V., Steinmetz, J. P., Blumen, H. M., Verghese, J., & Allali, G. (2016). Poor Gait Performance and Prediction of Dementia: Results From a Meta-Analysis. J Am Med Dir Assoc, 17(6), 482–490. 10.1016/j.jamda.2015.12.092

4. Bendayan, R., Cooper, R., Wloch, E. G., Hofer, S. M., Piccinin, A. M., & Muniz-Terrera, G. (2017). Hierarchy and Speed of Loss in Physical Functioning: A Comparison Across Older U.S. and English Men and Women. J Gerontol A Biol Sci Med Sci, 72(8), 1117–1122. 10.1093/gerona/glw209

5. Boutzoukas, E. M., O’Shea, A., Albizu, A., Evangelista, N. D., Hausman, H. K., Kraft, J. N., Van Etten, E. J., Bharadwaj, P. K., Smith, S. G., Song, H., Porges, E. C., Hishaw, A., DeKosky, S. T., Wu, S. S., Marsiske, M., Alexander, G. E., Cohen, R., & Woods, A. J. (2021). Frontal White Matter Hyperintensities and Executive Functioning Performance in Older Adults. Front Aging Neurosci, 13, 672535. 10.3389/fnagi.2021.672535

6. Carson, N., Leach, L., & Murphy, K. J. (2018). A re-examination of Montreal Cognitive Assessment (MoCA) cutoff scores. Int J Geriatr Psychiatry, 33(2), 379–388. 10.1002/gps.4756

7. Casaletto, K. B., Umlauf, A., Beaumont, J., Gershon, R., Slotkin, J., Akshoomoff, N., & Heaton, R. K. (2015). Demographically Corrected Normative Standards for the English Version of the NIH Toolbox Cognition Battery. J Int Neuropsychol Soc, 21(5), 378–391. 10.1017/S1355617715000351

8. Cecelja, M., McNally, R., Cleary, J., & Chowienczyk, P. (2025). Association Between Pulse Pressure and Structural Brain Changes in Aging: A Systematic Review and Meta-Analysis of Cross-Sectional and Longitudinal Studies. J Am Heart Assoc, 14(18), e040013. 10.1161/JAHA.124.040013

9. Clouston, S. A., Brewster, P., Kuh, D., Richards, M., Cooper, R., Hardy, R., Rubin, M. S., & Hofer, S. M. (2013). The dynamic relationship between physical function and cognition in longitudinal aging cohorts. Epidemiol Rev, 35(1), 33–50. 10.1093/epirev/mxs004

10. Cooper, R., Kuh, D., Hardy, R., Mortality Review, G., Falcon, & Teams, H. A. S. (2010). Objectively measured physical capability levels and mortality: systematic review and meta-analysis. BMJ, 341, c4467. 10.1136/bmj.c4467

11. Daimiel, L., Martinez-Gonzalez, M. A., Corella, D., Salas-Salvado, J., Schroder, H., Vioque, J., Romaguera, D., Martinez, J. A., Warnberg, J., Lopez-Miranda, J., Estruch, R., Cano-Ibanez, N., Alonso-Gomez, A., Tur, J. A., Tinahones, F. J., Serra-Majem, L., Mico-Perez, R. M., Lapetra, J., Galdon, A.,…Ordovas, J. M. (2020). Physical fitness and physical activity association with cognitive function and quality of life: baseline cross-sectional analysis of the PREDIMED-Plus trial. Sci Rep, 10(1), 3472. 10.1038/s41598-020-59458-6

12. de Jager, C. A., Budge, M. M., & Clarke, R. (2003). Utility of TICS-M for the assessment of cognitive function in older adults. Int J Geriatr Psychiatry, 18(4), 318–324. 10.1002/gps.830

13. de Oliveira Silva, F., Ferreira, J. V., Placido, J., Chagas, D., Praxedes, J., Guimaraes, C., Batista, L. A., Marinho, V., Laks, J., & Deslandes, A. C. (2019). Stages of mild cognitive impairment and Alzheimer’s disease can be differentiated by declines in timed up and go test: A systematic review and meta-analysis. Arch Gerontol Geriatr, 85, 103941. 10.1016/j.archger.2019.103941

14. Deary, I. J., Corley, J., Gow, A. J., Harris, S. E., Houlihan, L. M., Marioni, R. E., Penke, L., Rafnsson, S. B., & Starr, J. M. (2009). Age-associated cognitive decline. Br Med Bull, 92, 135–152. 10.1093/bmb/ldp033

15. Demnitz, N., Esser, P., Dawes, H., Valkanova, V., Johansen-Berg, H., Ebmeier, K. P., & Sexton, C. (2016). A systematic review and meta-analysis of cross-sectional studies examining the relationship between mobility and cognition in healthy older adults. Gait Posture, 50, 164–174. 10.1016/j.gaitpost.2016.08.028

16. Desjardins-Crepeau, L., Berryman, N., Vu, T. T., Villalpando, J. M., Kergoat, M. J., Li, K. Z., Bosquet, L., & Bherer, L. (2014). Physical functioning is associated with processing speed and executive functions in community-dwelling older adults. J Gerontol B Psychol Sci Soc Sci, 69(6), 837–844. 10.1093/geronb/gbu036

17. Diamond, A. (2013). Executive functions. Annu Rev Psychol, 64, 135–168. 10.1146/annurev-psych-113011-143750

18. Dias, J. M. (2014). Physical Functioning (PF). In A. C. Michalos (Ed.), Encyclopedia of quality of life and well-being research (1st 2014. ed., Vol. 1, pp. 4793–4795). Springer Reference.

19. Donoghue, O. A., Horgan, N. F., Savva, G. M., Cronin, H., O’Regan, C., & Kenny, R. A. (2012). Association between timed up-and-go and memory, executive function, and processing speed. J Am Geriatr Soc, 60(9), 1681–1686. 10.1111/j.1532-5415.2012.04120.x

20. Dourado, V. Z., Nishiaka, R. K., Simoes, M., Lauria, V. T., Tanni, S. E., Godoy, I., Gagliardi, A. R. T., Romiti, M., & Arantes, R. L. (2021). Classification of cardiorespiratory fitness using the six-minute walk test in adults: Comparison with cardiopulmonary exercise testing. Pulmonology, 27(6), 500–508. 10.1016/j.pulmoe.2021.03.006

21. Erickson, K. I., Leckie, R. L., & Weinstein, A. M. (2014). Physical activity, fitness, and gray matter volume. Neurobiol Aging, 35 *Suppl 2*, S20–28. 10.1016/j.neurobiolaging.2014.03.034

22. Erickson, K. I., Prakash, R. S., Voss, M. W., Chaddock, L., Hu, L., Morris, K. S., White, S. M., Wojcicki, T. R., McAuley, E., & Kramer, A. F. (2009). Aerobic fitness is associated with hippocampal volume in elderly humans. Hippocampus, 19(10), 1030–1039. 10.1002/hipo.20547

23. Ezzati, A., Katz, M. J., Lipton, M. L., Lipton, R. B., & Verghese, J. (2015). The association of brain structure with gait velocity in older adults: a quantitative volumetric analysis of brain MRI. Neuroradiology, 57(8), 851–861. 10.1007/s00234-015-1536-2

24. Fusco, O., Ferrini, A., Santoro, M., Lo Monaco, M. R., Gambassi, G., & Cesari, M. (2012). Physical function and perceived quality of life in older persons. Aging Clin Exp Res, 24(1), 68–73. 10.1007/BF03325356

25. Garcia Meneguci, C. A., Meneguci, J., Sasaki, J. E., Tribess, S., & Junior, J. S. V. (2021). Physical activity, sedentary behavior and functionality in older adults: A cross-sectional path analysis. PLoS One, 16(1), e0246275. 10.1371/journal.pone.0246275

26. Goodpaster, B. H., Park, S. W., Harris, T. B., Kritchevsky, S. B., Nevitt, M., Schwartz, A. V., Simonsick, E. M., Tylavsky, F. A., Visser, M., & Newman, A. B. (2006). The loss of skeletal muscle strength, mass, and quality in older adults: the health, aging and body composition study. J Gerontol A Biol Sci Med Sci, 61(10), 1059–1064. 10.1093/gerona/61.10.1059

27. Graham, J. E., Ostir, G. V., Fisher, S. R., & Ottenbacher, K. J. (2008). Assessing walking speed in clinical research: a systematic review. J Eval Clin Pract, 14(4), 552–562. 10.1111/j.1365-2753.2007.00917.x

28. Gu, Q., Zou, L., Loprinzi, P. D., Quan, M., & Huang, T. (2019). Effects of Open Versus Closed Skill Exercise on Cognitive Function: A Systematic Review. Front Psychol, 10, 1707. 10.3389/fpsyg.2019.01707

29. Harada, C. N., Natelson Love, M. C., & Triebel, K. L. (2013). Normal cognitive aging. Clin Geriatr Med, 29(4), 737–752. 10.1016/j.cger.2013.07.002

30. Hartley, P., Monaghan, A., Donoghue, O. A., Kenny, R. A., & Romero-Ortuno, R. (2022). Exploring bi-directional temporal associations between timed-up-and-go and cognitive domains in the Irish longitudinal study on ageing (TILDA). Arch Gerontol Geriatr, 99, 104611. 10.1016/j.archger.2021.104611

31. Hedden, T., & Gabrieli, J. D. (2004). Insights into the ageing mind: a view from cognitive neuroscience. Nat Rev Neurosci, 5(2), 87–96. 10.1038/nrn1323

32. Hoekstra, T., Rojer, A. G. M., van Schoor, N. M., Maier, A. B., & Pijnappels, M. (2020). Distinct Trajectories of Individual Physical Performance Measures Across 9 Years in 60- to 70-Year-Old Adults. J Gerontol A Biol Sci Med Sci, 75(10), 1951–1959. 10.1093/gerona/glaa045

33. Inoue, Y., Shue, F., Bu, G., & Kanekiyo, T. (2023). Pathophysiology and probable etiology of cerebral small vessel disease in vascular dementia and Alzheimer’s disease. Mol Neurodegener, 18(1), 46. 10.1186/s13024-023-00640-5

34. Jackson, A. S., Sui, X., Hebert, J. R., Church, T. S., & Blair, S. N. (2009). Role of lifestyle and aging on the longitudinal change in cardiorespiratory fitness. Arch Intern Med, 169(19), 1781–1787. 10.1001/archinternmed.2009.312

35. Jones, C. J., Rikli, R. E., & Beam, W. C. (1999). A 30-s chair-stand test as a measure of lower body strength in community-residing older adults. Res Q Exerc Sport, 70(2), 113–119. 10.1080/02701367.1999.10608028

36. Kaminsky, L. A., American College of Sports Medicine., & American College of Sports Medicine. (2014). ACSM’s health-related physical fitness assessment manual (4th ed.). Wolters Kluwer Health/Lippincott Williams & Wilkins.

37. Kaminsky, L. A., Arena, R., Ellingsen, O., Harber, M. P., Myers, J., Ozemek, C., & Ross, R. (2019). Cardiorespiratory fitness and cardiovascular disease - The past, present, and future. Prog Cardiovasc Dis, 62(2), 86–93. 10.1016/j.pcad.2019.01.002

38. Levine, D. A., Gross, A. L., Briceno, E. M., Tilton, N., Giordani, B. J., Sussman, J. B., Hayward, R. A., Burke, J. F., Hingtgen, S., Elkind, M. S. V., Manly, J. J., Gottesman, R. F., Gaskin, D. J., Sidney, S., Sacco, R. L., Tom, S. E., Wright, C. B., Yaffe, K., & Galecki, A. T. (2021). Sex Differences in Cognitive Decline Among US Adults. JAMA Netw Open, 4(2), e210169. 10.1001/jamanetworkopen.2021.0169

39. Lundberg, K., Elmstahl, S., Wranker, L. S., & Ekstrom, H. (2024). The Association between Physical Frailty and Cognitive Performance in Older Adults Aged 60 to 96 Years: Data from the "Good Aging in Skane" (GAS) Swedish Population Study. Ann Geriatr Med Res, 28(3), 330–341. 10.4235/agmr.24.0055

40. Lynch, D. H., Spangler, H., Griffin, J. S., Kahkoska, A., Boccaccio, D., Xie, W., Lin, F.-C., Batsis, J. A., & Fielding, R. A. (2025). Identifying Older Adults at Risk of Accelerated Decline in Gait Speed and Grip Strength: Insights from the National Health and Aging Trends Study (NHATS). Journal of Ageing and Longevity, 5(2). 10.3390/jal5020019

41. Martin, K. L., Blizzard, L., Wood, A. G., Srikanth, V., Thomson, R., Sanders, L. M., & Callisaya, M. L. (2013). Cognitive function, gait, and gait variability in older people: a population-based study. J Gerontol A Biol Sci Med Sci, 68(6), 726–732. 10.1093/gerona/gls224

42. Milani, S. A., Marsiske, M., Cottler, L. B., Chen, X., & Striley, C. W. (2018). Optimal cutoffs for the Montreal Cognitive Assessment vary by race and ethnicity. Alzheimers Dement (Amst), 10, 773–781. 10.1016/j.dadm.2018.09.003

43. Murray, M. E., Senjem, M. L., Petersen, R. C., Hollman, J. H., Preboske, G. M., Weigand, S. D., Knopman, D. S., Ferman, T. J., Dickson, D. W., & Jack, C. R., Jr. (2010). Functional impact of white matter hyperintensities in cognitively normal elderly subjects. Arch Neurol, 67(11), 1379–1385. 10.1001/archneurol.2010.280

44. Nasreddine, Z. S., Phillips, N. A., Bedirian, V., Charbonneau, S., Whitehead, V., Collin, I., Cummings, J. L., & Chertkow, H. (2005). The Montreal Cognitive Assessment, MoCA: a brief screening tool for mild cognitive impairment. J Am Geriatr Soc, 53(4), 695–699. 10.1111/j.1532-5415.2005.53221.x

45. Noble, C., Medin, D., Quail, Z., Young, C., & Carter, M. (2021). How Does Participation in Formal Education or Learning for Older People Affect Wellbeing and Cognition? A Systematic Literature Review and Meta-Analysis. Gerontol Geriatr Med, 7, 2333721420986027. 10.1177/2333721420986027

46. Oberlin, L. E., Wan, L., Kang, C., Romano, A., Aghjayan, S., Lesnovskaya, A., Ripperger, H. S., Drake, J., Harrison, R., Collins, A. M., Molina-Hidalgo, C., Grove, G., Huang, H., Kramer, A., Hillman, C. H., Burns, J. M., Vidoni, E. D., McAuley, E., Kamboh, M. I.,…Erickson, K. I. (2025). Cardiorespiratory fitness is associated with cognitive function in late adulthood: baseline findings from the IGNITE study. Br J Sports Med, 59(3), 167–176. 10.1136/bjsports-2024-108257

47. Ogawa, S., Himuro, N., Koyama, M., Seko, T., Mori, M., & Ohnishi, H. (2022). Walking Speed Is Better Than Hand Grip Strength as an Indicator of Early Decline in Physical Function with Age in Japanese Women Over 65: A Longitudinal Analysis of the Tanno-Sobetsu Study Using Linear Mixed-Effects Models. Int J Environ Res Public Health, 19(23). 10.3390/ijerph192315769

48. Pase, M. P., Herbert, A., Grima, N. A., Pipingas, A., & O’Rourke, M. F. (2012). Arterial stiffness as a cause of cognitive decline and dementia: a systematic review and meta-analysis. Intern Med J, 42(7), 808–815. 10.1111/j.1445-5994.2011.02645.x

49. Prasad, L., Fredrick, J., & Aruna, R. (2021). The relationship between physical performance and quality of life and the level of physical activity among the elderly. J Educ Health Promot, 10, 68. 10.4103/jehp.jehp_421_20

50. Prokopidis, K., Giannos, P., Ispoglou, T., Kirk, B., Witard, O. C., Dionyssiotis, Y., Scott, D., Macpherson, H., Duque, G., & Isanejad, M. (2023). Handgrip strength is associated with learning and verbal fluency in older men without dementia: insights from the NHANES. Geroscience, 45(2), 1049–1058. 10.1007/s11357-022-00703-3

51. Qu, Y., Hu, H. Y., Ou, Y. N., Shen, X. N., Xu, W., Wang, Z. T., Dong, Q., Tan, L., & Yu, J. T. (2020). Association of body mass index with risk of cognitive impairment and dementia: A systematic review and meta-analysis of prospective studies. Neurosci Biobehav Rev, 115, 189–198. 10.1016/j.neubiorev.2020.05.012

52. Reijnierse, E. M., de Jong, N., Trappenburg, M. C., Blauw, G. J., Butler-Browne, G., Gapeyeva, H., Hogrel, J. Y., McPhee, J. S., Narici, M. V., Sipila, S., Stenroth, L., van Lummel, R. C., Pijnappels, M., Meskers, C. G. M., & Maier, A. B. (2017). Assessment of maximal handgrip strength: how many attempts are needed? J Cachexia Sarcopenia Muscle, 8(3), 466–474. 10.1002/jcsm.12181

53. Rossetti, H. C., Lacritz, L. H., Cullum, C. M., & Weiner, M. F. (2011). Normative data for the Montreal Cognitive Assessment (MoCA) in a population-based sample. Neurology, 77(13), 1272–1275. 10.1212/WNL.0b013e318230208a

54. Rosso, A. L., Studenski, S. A., Chen, W. G., Aizenstein, H. J., Alexander, N. B., Bennett, D. A., Black, S. E., Camicioli, R., Carlson, M. C., Ferrucci, L., Guralnik, J. M., Hausdorff, J. M., Kaye, J., Launer, L. J., Lipsitz, L. A., Verghese, J., & Rosano, C. (2013). Aging, the central nervous system, and mobility. J Gerontol A Biol Sci Med Sci, 68(11), 1379–1386. 10.1093/gerona/glt089

55. Rosso, A. L., Verghese, J., Metti, A. L., Boudreau, R. M., Aizenstein, H. J., Kritchevsky, S., Harris, T., Yaffe, K., Satterfield, S., Studenski, S., & Rosano, C. (2017). Slowing gait and risk for cognitive impairment: The hippocampus as a shared neural substrate. Neurology, 89(4), 336–342. 10.1212/WNL.0000000000004153

56. Singh, B., Bennett, H., Miatke, A., Dumuid, D., Curtis, R., Ferguson, T., Brinsley, J., Szeto, K., Petersen, J. M., Gough, C., Eglitis, E., Simpson, C. E., Ekegren, C. L., Smith, A. E., Erickson, K. I., & Maher, C. (2025). Effectiveness of exercise for improving cognition, memory and executive function: a systematic umbrella review and meta-meta-analysis. Br J Sports Med, 59(12), 866–876. 10.1136/bjsports-2024-108589

57. Song, J., Lindquist, L. A., Chang, R. W., Semanik, P. A., Ehrlich-Jones, L. S., Lee, J., Sohn, M. W., & Dunlop, D. D. (2015). Sedentary Behavior as a Risk Factor for Physical Frailty Independent of Moderate Activity: Results From the Osteoarthritis Initiative. Am J Public Health, 105(7), 1439–1445. 10.2105/AJPH.2014.302540

58. Su, Y. H., Chiou, J. M., Shiu, C., Chen, J. H., & Chen, Y. C. (2025). Longitudinal, Bidirectional Association between Gait Speed and Cognitive Function in Community-Dwelling Older Adults without Dementia. J Am Med Dir Assoc, 26(5), 105544. 10.1016/j.jamda.2025.105544

59. Thomas, S., Reading, J., & Shephard, R. J. (1992). Revision of the Physical Activity Readiness Questionnaire (PAR-Q). Can J Sport Sci, 17(4), 338–345.

60. Tian, Q., Chastan, N., Bair, W. N., Resnick, S. M., Ferrucci, L., & Studenski, S. A. (2017). The brain map of gait variability in aging, cognitive impairment and dementia-A systematic review. Neurosci Biobehav Rev, 74(Pt A), 149–162. 10.1016/j.neubiorev.2017.01.020

61. Tromp, D., Dufour, A., Lithfous, S., Pebayle, T., & Despres, O. (2015). Episodic memory in normal aging and Alzheimer disease: Insights from imaging and behavioral studies. Ageing Res Rev, 24(Pt B), 232–262. 10.1016/j.arr.2015.08.006

62. Tulving, E. (2002). Episodic memory: from mind to brain. Annu Rev Psychol, 53, 1–25. 10.1146/annurev.psych.53.100901.135114

63. United Nations Department of Economic and Social Affairs. (2023). *World Social Report 2023: Leaving No One Behind in an Ageing World* (9789211304589). https://digitallibrary.un.org/record/4000104/files/2023wsr-fullreport.pdf

64. Van Dam, N. T., & Earleywine, M. (2011). Validation of the Center for Epidemiologic Studies Depression Scale--Revised (CESD-R): pragmatic depression assessment in the general population. Psychiatry Res, 186(1), 128–132. 10.1016/j.psychres.2010.08.018

65. van Iersel, M. B., Munneke, M., Esselink, R. A., Benraad, C. E., & Olde Rikkert, M. G. (2008). Gait velocity and the Timed-Up-and-Go test were sensitive to changes in mobility in frail elderly patients. J Clin Epidemiol, 61(2), 186–191. 10.1016/j.jclinepi.2007.04.016

66. Verhaeghen, P., & Salthouse, T. A. (1997). Meta-analyses of age-cognition relations in adulthood: estimates of linear and nonlinear age effects and structural models. Psychol Bull, 122(3), 231–249. 10.1037/0033-2909.122.3.231

67. Wang, J., Cui, Q., Xu, X., & Yang, G. (2025). Bidirectional association between grip strength and cognitive function in Chinese older adults: a nationwide cohort study. BMC Public Health, 25(1), 1880. 10.1186/s12889-025-23079-3

68. Weintraub, S., Dikmen, S. S., Heaton, R. K., Tulsky, D. S., Zelazo, P. D., Bauer, P. J., Carlozzi, N. E., Slotkin, J., Blitz, D., Wallner-Allen, K., Fox, N. A., Beaumont, J. L., Mungas, D., Nowinski, C. J., Richler, J., Deocampo, J. A., Anderson, J. E., Manly, J. J., Borosh, B.,…Gershon, R. C. (2013). Cognition assessment using the NIH Toolbox. Neurology, 80(11 Suppl 3), S54–64. 10.1212/WNL.0b013e3182872ded

69. Wollesen, B., Wildbredt, A., van Schooten, K. S., Lim, M. L., & Delbaere, K. (2020). The effects of cognitive-motor training interventions on executive functions in older people: a systematic review and meta-analysis. Eur Rev Aging Phys Act, 17, 9. 10.1186/s11556-020-00240-y

70. Yan, S., Fu, W., Wang, C., Mao, J., Liu, B., Zou, L., & Lv, C. (2020). Association between sedentary behavior and the risk of dementia: a systematic review and meta-analysis. Transl Psychiatry, 10(1), 112. 10.1038/s41398-020-0799-5

71. Yang, E. J., Lim, S., Lim, J. Y., Kim, K. W., Jang, H. C., & Paik, N. J. (2012). Association between muscle strength and metabolic syndrome in older Korean men and women: the Korean Longitudinal Study on Health and Aging. Metabolism, 61(3), 317–324. 10.1016/j.metabol.2011.07.005

72. Yogev-Seligmann, G., Hausdorff, J. M., & Giladi, N. (2008). The role of executive function and attention in gait. Mov Disord, 23(3), 329–342; quiz 472. 10.1002/mds.21720

73. Yoon, D. H., Hwang, S. S., Lee, D. W., Lee, C. G., & Song, W. (2018). Physical Frailty and Cognitive Functioning in Korea Rural Community-Dwelling Older Adults. J Clin Med, 7(11). 10.3390/jcm7110405

74. Zammit, A. R., Robitaille, A., Piccinin, A. M., Muniz-Terrera, G., & Hofer, S. M. (2019). Associations Between Aging-Related Changes in Grip Strength and Cognitive Function in Older Adults: A Systematic Review. J Gerontol A Biol Sci Med Sci, 74(4), 519–527. 10.1093/gerona/gly046

